# Machine Learning Models for Diagnosis of Cushing’s Syndrome Using Retrospective Data

**DOI:** 10.1101/2020.05.31.20118604

**Authors:** Senol Isci, Derya Sema Yaman Kalender, Firat Bayraktar, Alper Yaman

## Abstract

Accurate classification of Cushing’s Syndrome (CS) plays a critical role in providing early and correct diagnosis of CS that may facilitate treatment and improve patient outcomes. Diagnosis of CS is a complex process, which requires careful and concurrent interpretation of signs and symptoms, multiple biochemical test results, and findings of medical imaging by physicians with a high degree of specialty and knowledge to make correct judgments. In this article, we explore the state of the art machine learning algorithms to demonstrate their potential as a clinical decision support system to analyze and classify CS in order to facilitate the diagnosis, prognosis, and treatment of CS. Prominent algorithms are compared using nested cross-validation and various class comparison strategies including multiclass, one vs. all, and one vs. one binary classification. Our findings show that Random Forest (RF) algorithm is most suitable for the classification of CS. We demonstrate that the proposed approach can classify CS subjects with an average accuracy of 92% and an average F1 score of 91.5%, depending on the class comparison strategy and selected features. RF-based one vs. all binary classification model achieves sensitivity of 97.6%, precision of 91.1%, and specificity of 87.1% to discriminate CS from non-CS on the test dataset. RF-based multiclass classification model achieves average per class sensitivity of 91.8%, average per class specificity of 97.1%, and average per class precision of 92.1% to classify different subtypes of CS on the test dataset. Clinical performance evaluation suggests that the developed models can help improve physician’s judgment in diagnosing CS.

## 1. Introduction

Cushing’s Syndrome (CS) is a potentially lethal disorder caused by abnormally high levels of cortisol hormone, first described in 1912 by Harvey Cushing [1],[2]. Early diagnosis plays a crucial role in reducing mortality and improving the prognosis of this syndrome [3]. However, the diagnosis of CS can be difficult, for instance, due to gradual development of symptoms, and due to overlap with features of metabolic syndrome like increased blood pressure, high blood sugar, excess body fat around the waist, and abnormal cholesterol or triglyceride levels. Moreover, many of these features are common in the general population [3].

Diagnosis of CS is a complex process, which requires careful and concurrent interpretation of signs and symptoms, results of multiple biochemical test measurements, and findings of medical imaging by physicians with a high degree of specialty and knowledge to make correct decisions. Variations in accuracy of biochemical tests, differences in test and decision criteria, and overlapping characteristics of CS subtypes make it even harder. In this study, we show that machine learning (ML) models that utilize selected input features derived from biochemical tests and imaging findings of patients who are suspected of CS can classify nonfunctional adrenal adenoma, subclinical CS, adrenal CS, and pituitary CS at screening test stage and follow-up test stage for the diagnosis of CS.

### 1.1. Background

Excessive level of cortisol (i.e., hypercortisolism) shows clinical characteristics like a large rounded face, abdominal obesity, and diminishing of muscle tissue or skin thickness. Early diagnosis plays a crucial role in reducing mortality and improving the prognosis of this syndrome [3]. However, the diagnosis of CS can be difficult, for instance, due to gradual development of symptoms, and due to overlap with features of metabolic syndrome like increased blood pressure, high blood sugar, excess body fat around the waist, and abnormal cholesterol or triglyceride levels. Moreover, many of these features are common in the general population [3]. The estimated incidence of CS is 0.2–5 per 1 million per year, and its prevalence is 39–79 per million in various populations. The median age is 41.4 years, and the female to male ratio is 4 to 1 [4], [5].

There are several causes of CS. It may stem from prolonged intake of glucocorticoids-steroid hormones that are chemically similar to natural cortisol, such as anti-inflammatory medications prescribed for asthma, rheumatoid arthritis, lupus, and other inflammatory diseases. Such hormones may also be taken after an organ transplant to suppress the immune system and prevent organ rejection. There are also endogenic causes in which the body produces an excessive amount of cortisol by itself. Cushing Disease, a form of CS is the most common cause of excess endogenous cortisol production by the adrenal glands. It is caused by a pituitary tumor (i.e., adenoma which is usually a benign tumor in glands) that secretes excessive amount of adrenocorticotropic hormone (ACTH), which then signals the adrenal glands to produce cortisol. CS might be a result of an adrenal gland tumor or adrenal hyperplasia (i.e., a genetic disorder in the adrenal gland), which can cause the adrenal gland to overproduce cortisol. Ectopic CS is another form of CS in which a tumor in another part of the body such as the pancreas, lung, or thyroid can result in CS by producing ACTH. It is called ectopic ACTH production because it is produced somewhere other than the pituitary gland.

Laboratory investigations for patients with clinically suspected CS are divided into two stages. Stage-1 tests are screening tests for diagnostic purposes and applied to prove the presence of hypercortisolism. Stage-2 includes follow up tests to evaluate the cause of hypercortisolism [5], [6], [7]. The most commonly used evaluation procedure for the exclusion or confirmation of CS is urine cortisol test which measures the cortisol level in a 24-hour sample of urine, saliva cortisol test which measures the cortisol level in the saliva, and low-dose dexamethasone test which measures the cortisol level in the blood after intake of the drug Dexamethasone. Despite the predictive value of these methods, cases with inconclusive test results still happen. Inconclusive results can be seen in patients with initial stages of this disease or in periodic forms of CS. Sometimes the diagnosis can only be made after long-term follow up or prolonged procedures, and may require hospitalization of the patient [6]. For patients with incidental adrenal mass, it is difficult to make a diagnosis or operation decision only with test results [8]. Incidental adrenal mass refers to the incidentally discovered adrenal mass during imaging which was not performed for suspected adrenal disease. Some conditions (e.g., obesity, diabetes mellitus or depression) which have common features with CS may cause physiological hypercortisolism, and lead to incorrect results of the Dexamethasone suppression test (DST) [9].

### 1.2. Motivations

There are several difficulties in diagnosing CS. Factors like laboratory errors, patient-induced errors, differences between groups, age, and gender may cause inconsistent test results. Traditionally, the diagnostic values of the test results have been analyzed using statistical methods and based on different selected cut-offs for sensitivity and specificity tradeoff. Therefore, evaluation of the tests performed for the diagnosis and the identification of the cause may significantly vary. In many studies, different cut-off values have been found depending on the varying settings of the medical test and employed statistical methods [10]. The diagnosis of CS and identification of its cause require careful and concurrent interpretation of signs and symptoms, results of multiple biochemical test measurements, and findings of medical imaging by physicians with a high degree of specialty and knowledge to make correct judgments.

We believe that these difficulties would be handled by utilizing a generalizable machine learning (ML) approach. In this article, we explore the state of the art ML algorithms, and demonstrate their usefulness as a clinical decision support system to evaluate results of the medical tests, and predict CS in order to facilitate the diagnosis and prognosis of CS.

### 1.3. Contributions

The proposed approach is suitable for use in both screening testing stage for the diagnosis of CS, and follow-up testing stage for the identification of the cause of CS. Several prominent ML algorithms were compared using nested cross-validation (NCV) and using various class comparison strategies including multiclass, one vs. all, and one vs. one binary classifications in order to optimize model parameters and estimate algorithm/model performance in classification of CS types and non-CS, namely, pituitary CS, adrenal CS, subclinical CS, and nonfunctional adrenal adenoma. First, Support Vector Machines (SVM), K-nearest Neighbors, Logistic Regression, Linear Discriminant Analysis, Decision Tree (DT), Random Forest (RF), Adaptive Boosting, and Gradient Boosting were considered as classifier algorithms. According to the algorithm/model comparison and selection process, we found that RF algorithm is suitable for the classification of CS. Subsequently, RF-based models were developed using training and validation (T&V) dataset with selected features derived from measurements of Stage-1 biochemical screening test, Stage-2 biochemical follow-up tests, and imaging findings. Trained models were tested using separate independent dataset to show their predictive performance on unseen data in a real-world setting. The developed models were used for further analysis and classification study of CS. Furthermore, clinical performance evaluation of the suggested method was performed by comparing model predictions to the judgments of expert physicians.

We show that using ML approach, it is possible to classify subjects with CS with an average accuracy of 92% and an average F1 score of 91.5%, depending on the class comparison strategy and the selected features. RF-based one vs. all binary classification model achieves sensitivity of 97.6%, precision of 91.1%, and specificity of 87.1% to separate CS from nonfunctional adrenal adenoma on the final independent test (FIT) dataset. RF-based multiclass classification model achieves average per class sensitivity of 91.8%, average per class specificity of 97.1%, and average per class precision of 92.1% to classify types of CS on the FIT dataset. We suggest the use of binary classification model with features derived from Stage-1 tests to discriminate between nonfunctional adrenal adenoma and CS for the diagnosis of CS during clinical evaluations. We suggest the use of multiclass model with features derived from both Stage-1 and Stage-2 tests to discriminate among types of CS to find the cause of CS during clinical evaluations. The results support that the proposed approach can be valuable in the diagnosis, prognosis, and treatment of CS.

The developed ML models outperformed the physician’s judgments with the constraint of using only the selected biochemical test results utilized in ML model development. In Stage-1, physicians were able to classify subjects with F1 score of 76.6 % whereas F1 score of ML model was 98.5%. In Stage-2, the overall success rate of the physicians was low with an F1 score of 49.4% while F1 score of ML model was 96.4%.

We also developed a web application as clinical decision support tool for public use where a user can input patient’s test findings and receive prediction for CS.

To the best of our knowledge, this is the first study to present a comprehensive investigation of the application of ML approach to early detection and classification of CS using retrospective medical data.

The remainder of the paper is structured as follows. The materials and technical aspects of this work are outlined in Section 2. The experimental investigations are reported in Section 3 with results analyzed. The discussions about merits, limitations, justifications to selected methods and relevant further research is given in Section 4. The article concludes in Section 5.

## 2. Materials and Technical Approach

### 2.1. Materials

In this study, the medical records of 241 subjects were used, who were admitted to the endocrinology outpatient clinic of Dokuz Eylul University Medical Faculty due to CS symptoms or incidental adrenal adenomas between 2005 and 2016. The average age of the subjects was 52.02±13.33 years. 183 subjects were female whereas 58 of them were male. Out of 241 subjects, 104 subjects (55.24± 10.46 years, 71 female, 33 male) had nonfunctional adrenal adenomas, 59 subjects (56.88± 12.65 years, 47 female, 12 male) had subclinical CS; 42 subjects (47.90± 13.69 years, 33 female, 9 male) had adrenal CS, and 36 subjects (39.55± 12.68 years, 32 female, 4 male) had pituitary CS. We excluded 3 subjects of ectopic CS since data samples were not sufficient for further processing. The results of diagnostic tests including retrospective basal cortisol, basal ACTH, 1mg DST cortisol, 2mg DST cortisol, 8mg DST cortisol, midnight cortisol, 24-hour urine cortisol, and adrenal and pituitary imaging tests of the diagnosed and monitored subjects were examined. The normal reference range of urinary cortisol was 58–403 μg / 24h in the hospital. Dokuz Eylul University Faculty of Medicine Ethics Committee approved this research study and waived the requirement for consent (2019/28–26).

### 2.2. Overview of the overall approach

Fig. 1 shows a flowchart of the overall system. Input dataset (n = 241, see Table 1) was randomly split into T&V dataset (n = 168, 70%), and FIT dataset (n = 73, 30%) by stratified sampling ensuring that percentages of observations in each of 4 classes (i.e., NF, SC, AD, and PT) are preserved in the created datasets. Two-sample Kolmogorov-Smirnov test[11] was applied to check whether both of T&V and FIT datasets are representative of the same distribution. We used median imputation method to replace missing values in the T&V dataset with the median values calculated for each feature from the T&V dataset. Namely, the missing data for a given feature was replaced by the median of all known values of that feature. Similarly, medians were calculated from the FIT dataset and applied to missing values in the FIT dataset. Binary features were treated as continuous features. A number of the feature distributions were observed to be skewed. Therefore, logarithmic transformation was applied to the whole dataset to make skewed feature distributions less skewed, approximating to normal distribution. Transformation was performed after the imputation because it changes underlying distribution. Logarithmic transformation helps the range and variance of features to be on a comparable scale; therefore, no further scaling was performed. Datasets were created according to class comparison strategy including multiclass, one vs. one binary classification, and one vs. all binary classification to be used for training and evaluating ML algorithms in terms of their suitability to classify CS types and non-CS cases, namely, pituitary CS (PT), adrenal CS (AD), subclinical CS (SC), and nonfunctional adrenal adenoma (NF).

**Fig. 1.**
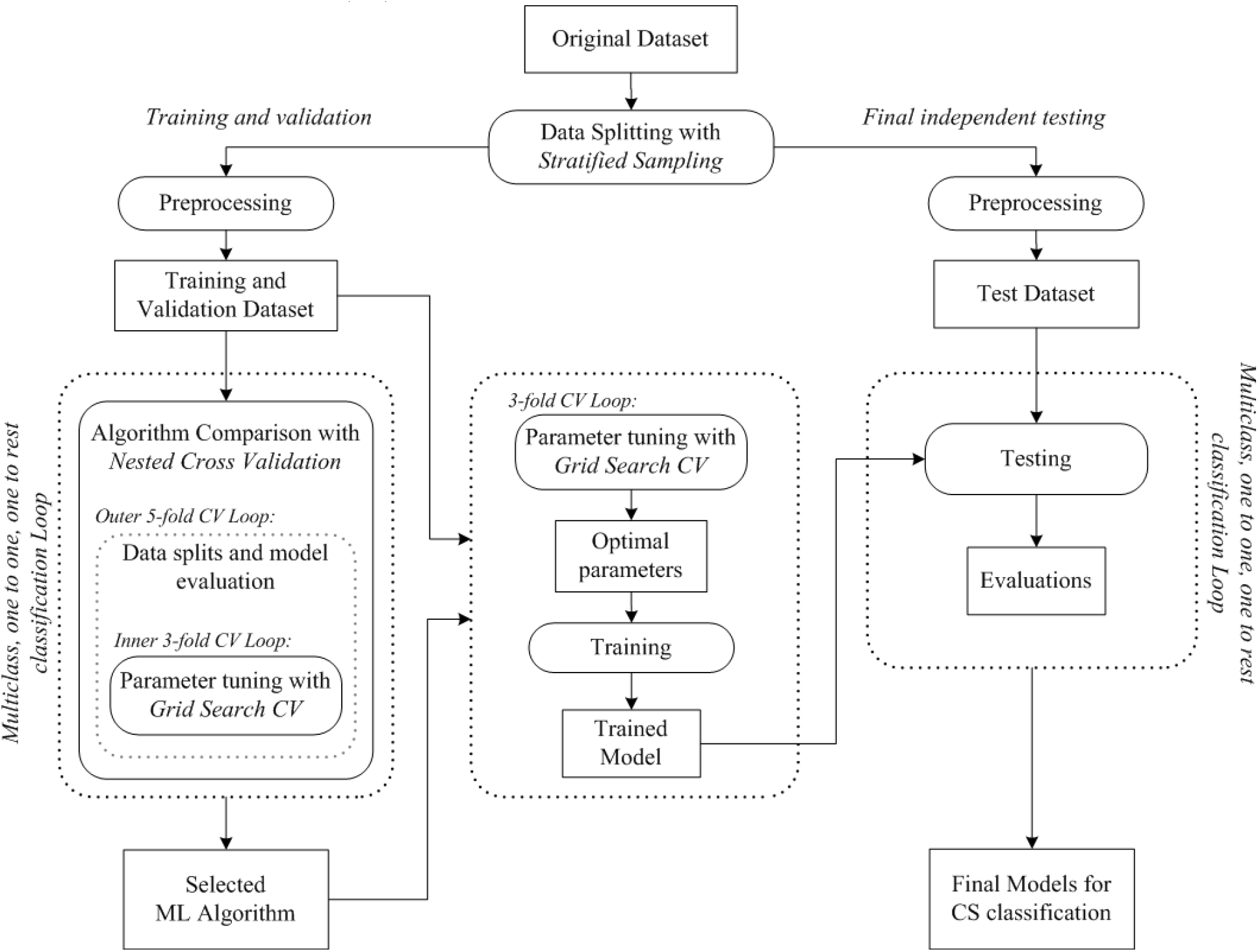
The flowchart of the overall method.

**Table 1.** Properties of the dataset.

| Feature | Description | Missing Value Ratio (%) | Diagnosis |  |  |  |
| --- | --- | --- | --- | --- | --- | --- |
|  |  |  | NF (n=104) | SC (n=59) | AD (n=42) | PT (n=36) |
| age | age of the subject (years) | 0% | 55.24 ± 10.46 | 56.88 ± 12.65 | 47.9 ± 13.7 | 39.56 ± 12.69 |
| gender | gender of the subject (female/male count) | 0% | 71/33 | 47/12 | 33/9 | 32/4 |
| bc | basal cortisol (µg/dL) | 2% | 13.84 ± 6.05 | 13.55 ± 4.48 | 20.64 ± 7.82 | 21.38 ± 10.9 |
| bacth | basal ACTH (pg/mL) | 2% | 20.88 ± 14.15 | 10.62 ± 7 | 9.09 ± 8.59 | 59.08 ± 46.89 |
| 1mgDSTc | cortisol after 1mg DST (µg/dL) | 11% | 1.37 ± 1.24 | 4.63 ± 4.22 | 13.3 ± 8.43 | 12.48 ± 5.7 |
| 2mgDSTc | cortisol after 2mg DST (µg/dL) | 49% | 1.77 ± 1.02 | 4.36 ± 3.47 | 14.58 ± 10.69 | 12.3 ± 7.33 |
| 8mgDSTc | cortisol after 8mg DST (µg/dL) | 71% | - | 5 ± 5.61 | 14.79 ± 10.99 | 5.15 ± 5.59 |
| mc | midnight cortisol (µg/dL) | 32% | 3.87 ± 2.63 | 6.43 ± 3.33 | 16.16 ± 6.71 | 18.69 ± 9.61 |
| ufc | urinary free cortisol (µg/24h)<br>hospital reference ufc range: 58-403 µg / 24h. | 33% | 156.32 ± 79.63 | 181.29 ± 157.71 | 307.04 ± 294.07 | 339.34 ± 286.12 |
| adrMass | mass in adrenal imaging (yes/no count) | 0% | 104/0 | 59/0 | 42/0 | 8/28 |
| pitMass | mass in pituitary imaging (yes/no count) | 0% | 1/103 | 8/51 | 4/38 | 36/0 |

We compared 8 different classification algorithms: Support Vector Machines, K-nearest Neighbors, Logistic Regression, Linear Discriminant Analysis, Decision Tree, Random Forest, Adaptive Boosting, and Gradient Boosting. Features were selected according to the physician’s expert knowledge and two-staged medical testing approach in clinical diagnosis of CS. Table 2 summarizes the class comparison strategy, associated data sizes, and selected feature sets for the diagnostic testing stages (i.e., Stage-1 and Stage-2). The estimates of predictive performances of the algorithms were assessed by nested cross-validation (NCV) approach. The 3 fold inner loop of NCV procedure was used to tune the hyperparameters of the created models using predefined possible values. The 5 fold outer loop of the NCV procedure was then used to assess the generalization performance of the models. The NCV procedure was repeated for all algorithms, class comparison strategies, and different set of features. The algorithm which achieves an overall total performance was selected, which turned out to be the RF algorithm in this study. After algorithm comparison and selection, the optimal hyperparameters of the best performing algorithm were selected in 3-fold cross-validation (CV) with a grid search procedure using the data folds from the complete T&V dataset. Grid search refers to the exhaustive search over specified hyperparameter values. The selected parameters were then used to train the classification model on the complete T&V dataset. The trained models were evaluated on the FIT dataset for their predictive performance for the case of unseen data. At the final step, the complete dataset was used to train final models to be used for te*s*ting future samples.

**Table 2.**
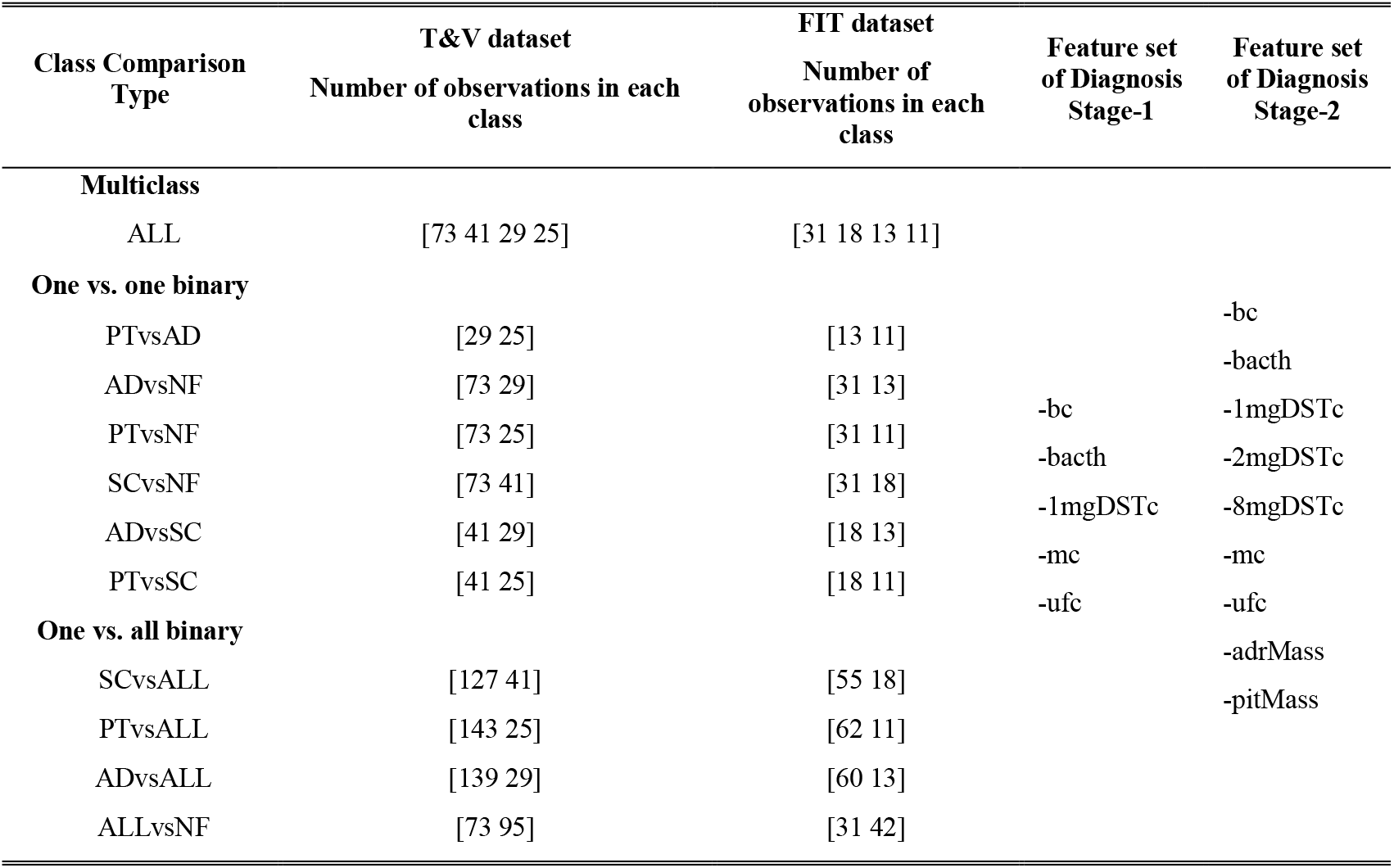
Class comparison strategies, associated datasets, and features.

We used the Python scikit-learn ML library v0.21.3 [12], which contains implementations of all models used in this study. Software code is available at https://github.com/SenolIsci/cushing01.

The web application of the decision support tool is publicly accessible at https://cushings-syndrome-prediction.herokuapp.com/.

The following sections provide the details of steps, procedures, and algorithms of the overall technical approach.

### 2.3. Dataset profile

The CS dataset consists of 241 samples, 11 features (i.e., predictor variables), and the target variable representing 4 classes for diagnostic type of CS. Dataset is considered to be a relatively limited dataset. The statistical properties (i.e., mean, standard deviation, ratio, and counts (n)) of the dataset features according to the type of diagnosis are given in Table 1.

For a better understanding of the structure of the dataset, we examined the feature distributions within the dataset. A number of the feature distributions were observed to be skewed (i.e., asymmetry of the distribution about its mean) especially all of the distributions of cortisol and ACTH related features (see Fig. S1 of Supplementary Materials).

Dataset has missing values because some of the medical tests were not performed depending on the symptoms and results of the accompanying tests as decided by the physicians. Dataset faces a moderate class imbalance problem because one of the classes (i.e., NF) is represented by a large number of samples whereas the others are represented by fewer samples. Features age and gender were reported for reference purposes, and they were not used for the rest of the study in order to focus only on features representing medical tests.

### 2.4. Dataset splitting and subset preparation

Input dataset (n = 241) was randomly split into T&V dataset (n = 168, 70%) and FIT dataset (n = 73, 30%) by stratified sampling ensuring that ratios of classes are represented in the newly created datasets. Due to the limited dataset size, two-sample Kolmogorov-Smirnov (K-S) test was performed to check whether both of T&V and FIT datasets are representative of the same distribution. A representative test dataset is necessary to provide sufficient information to evaluate the ability of the trained model to generalize to unseen data. The test was applied for each feature in the datasets. If the K-S statistics of the test is small or the p-value is high, then we cannot reject the hypothesis at the selected 5% significance level that the distributions of the two samples are similar. The procedure is tested for repetitiveness over 10 sampling cases and randomly chosen data subsets were used. In this study, two-sample Kolmogorov-Smirnov test yielded high values of p-values between 0.354 and 1 and K-S statistics between 0.003 and 0.128, indicating that the datasets were representative of the same distribution (see Table S1 of Supplementary Materials).

We intend to evaluate multiclass, one vs. all, and one vs. one binary classification strategies for the task of CS type classification. Therefore, data subsets were created accordingly. Multiclass strategy consists of fitting one classifier for all classes. One vs. one strategy consists of fitting one classifier per class pair. One vs. all strategy consists of fitting one classifier per class, and for each classifier, the class is fitted against all the other classes. The dataset properties according to class comparison strategy and feature sets in Stage-1 and Stage-2 diagnostics stages are listed in Table 2.

### 2.5. Dealing with missing values

The treatment of missing data is a broad statistical problem, and there is no universal imputation method performing best in every situation [13]. The simplest option is discarding samples with missing values. However, in this study, dropping samples with missing values of features was not considered as an option because missing values are ubiquities in the original dataset. This means that such omissions would result in severe information loss and a very small dataset. The reason for missing values of features is because some of the medical tests for a subject are not performed as a result of the medical testing and diagnosis procedures decided by the physicians, as mentioned in Section 1.1 and detailed in Section 1 of Supplementary Materials. For example, 8 mg DST test is performed for identifying the type of CS only after the screening tests such as 1mg DST. If a subject is believed to have nonfunctional adrenal adenoma, 8 mg DST is almost always skipped. Or depending on the severity of symptoms and other indicators, the physician may decide to skip low dose DST test and jump to 8 mg DST test. Therefore, we avoided omitting features with missing values and used imputation.

In this study, we separately applied median imputation on T&V dataset and FIT dataset. The missing value for a given feature was replaced by the median of all known values of that feature calculated separately both for T&V dataset and FIT dataset. In this way, information leakage is prevented, and independence of the FIT dataset is preserved from the rest of the training process, however, at the expense of degrading statistics.

Although there are a number of advanced imputation methods such as KNN, Maximum Likelihood, and Multivariate imputation by chained equations (MICE), these methods typically introduce additional parameters that require tuning, and hence, have the risk of overfitting and reduced generalizability, especially in case of small dataset [14], [15], [16]. Due to the limited size of the dataset, the missing value problem was mitigated by using median imputation, which does not require optimization of any parameters. Moreover, median imputation is suitable when the distribution of a given feature is skewed, which is true in our case. However, as we continue to collect more data, more advanced imputation methods could be utilized.

### 2.6. Dealing with class imbalance problem

Classes are moderately imbalanced in our dataset of limited size in which there are 104 NF, 59 SC, 42 AD, 36 PT samples. Since the probability of observations belonging to the majority class is significantly high in imbalanced data set, the algorithms are much more likely to classify new observations to the majority class.

There are downsampling and several oversampling techniques that try to balance the dataset by increasing the size of rare samples for example by using repetition, bootstrapping, or generating synthetic data [17]. However, when the dataset is relatively small, downsampling the majority class leads to a significant reduction in dataset size and loss of information. Similarly, the most severe weakness of oversampling is that it adds no new information to the dataset, and it could cause overfitting of classifiers [17]. Another method to deal with the class imbalance problem is choosing the right ML algorithm that can inherently handle imbalanced data classification such as thanks to the partitioning mechanism in tree-based algorithms in an ensemble setting or capability of rebalancing the dataset by putting different weights on the majority class and minority class observations. Furthermore, in this study, we aim to compare algorithms for a real-world scenario where imbalance in the number of cases with different CS subtype is highly possible. Therefore, no oversampling or synthetic data generation method was employed, but the set of algorithms were selected accordingly, and their parameters were tuned for handling class imbalance problem. Also, suitable performance metrics were used for measuring model performance in case of imbalanced data.

### 2.7. Dealing with skewed feature distributions

Skewness is a measure of the degree of asymmetry of a distribution, in which the histogram appears distorted or skewed either to the left or right. If the left tail is more pronounced than the right tail, it is said to have negative skewness. If the reverse is true, it has positive skewness. If the two are equal, it has zero skewness. It can be measured by using Fisher-Pearson coefficient of skewness which is (3x(Mean-Median)/ Standard Deviation) [18], [19]. In a normal distribution, the histogram appears as a symmetrical “bell-shaped curve” (i.e., zero skewness). The mean, or average, and the mode, or maximum point on the curve, are equal.

The real datasets are generally found to be asymmetric or specifically skewed. Some ML algorithms such as LDA classifier have normality assumption for the underlying populations, and their performance is adversely affected by the violation of this assumption. This problem was demonstrated in a study to compare classifier performance on skewed data [20]. In general, skewed distributions of the feature in the dataset will degrade the model’s ability to describe more prevalent cases in order to deal with much rarer cases which happen to take extreme values. Some precision may be lost with the cases one observes frequently in order to accommodate cases that are less observed.

Fixing skewness can provide a variety of benefits, including making analysis which assumes normal feature distributions possible or more informative. It may also provide a comparable scale for feature values and prevent extreme values of the feature from overestimating or underestimating the influence of the skewed feature on the target classification.

Classifiers that are free of any distributional assumptions are expected to perform well with a variety of distributions as long as the class distributions are reasonably distinct. For example, RF algorithm is more tolerable to skewed feature distributions [21].

A transformation on feature distributions may be used to reduce skewness. One simple but powerful transformation is the logarithmic transformation (e.g., log base 10, or natural logarithm) which has a major effect on distribution shape. Logarithmic transformation aims to map data from any distribution to as close to a normal distribution as possible in order to stabilize variance and minimize skewness. It is often appropriate for continuous variables, and cannot be applied to distribution with zero or negative values.

In this study, we applied logarithmic transformation (log base 10) to the dataset in an effort to make feature distributions less skewed, and reported the level of skewness using Fisher-Pearson coefficient of skewness. Mean absolute skewness was reduced from 2.398 to 0.897 after transformation of the T&V dataset. The results of the skewness test before and after preprocessing of the dataset are given in Table S2 of Supplementary Materials.

### 2.8. Machine Learning Algorithms

We compared several ML algorithms, all of which can be employed in multiclass, one vs. all, and one vs. one binary classification strategy for the task of classification of CS. Multiclass strategy encompasses fitting of one classifier for all classes. One vs. one strategy includes fitting one classifier per class pair. One vs. all strategy consists of fitting one classifier per class, assuming the label of the class as positive and labels of other classes as negative. The algorithms evaluated for CS are as the following:

#### 2.8.1. Support Vector Machine

Support Vector Machine (SVM) discriminates a set of high-dimension features using hyperplanes that separate data belonging to different classes while maximizing the margin, or the distance of the training samples to the separating hyperplane [22]. In other words, we plot each data item as a point in n-dimensional space (where n is the number of features) with the value of each feature being the value of a particular coordinate. Then, we perform classification by finding the hyper-plane that best segregate the two classes. The decision function is fully specified by a usually very small subset of training samples, the support vectors. Support vectors are the data points that lie closest to the hyperplane. Kernel functions are introduced for non-linear separation problems in which classes are not linearly separable. These are functions that take low dimensional input space and transform it into a higher-dimensional space based on the output classes in the data.

#### 2.8.2. K-nearest Neighbor

K-nearest Neighbor (KNN) classifier is one of the basic ML algorithms for classification [23]. KNN is a type of instance-based learning or non-generalizing learning, which does not attempt to construct a general internal model but simply stores instances of the training data. The principle of this method is based on the idea that samples of the same class should be closer in the feature space. As a result, for a given data point of unknown class, the algorithm can compute the distance between the given data point of unknown class and all the data points in the training data, and assign the class determined by majority vote of the K nearest neighbor points of the given data point. The distance can be in general any metric measure: Euclidean distance is the most common choice. Tree-based algorithms have been developed to reduce the number of distance calculations such as K-D Tree [24] and Ball Tree [25].

#### 2.8.3. Logistic Regression

Logistic Regression (LG) classifier is a widely used method for modeling binary outcomes [26]. LG employs the logistic function, which is an S-shaped curve that can take any real-valued number and map it into a value between 0 and 1. The probabilities describing the possible outcomes are calculated using a logistic function. LG classifier algorithm uses an equation in which input values of variables are combined linearly using weights/coefficient values to predict an output value of the output variable (i.e., class label). The coefficients of the LG algorithm are estimated from the training data. This is done, for example, using maximum-likelihood estimation. Multinomial logistic regression generalizes logistic regression to multiclass problems, i.e. with more than two possible discrete outcomes.

#### 2.8.4. Linear Discriminant Analysis

Linear Discriminant Analysis Classifier is a classifier with a linear decision boundary, generated by fitting class conditional densities to the data, and by using Bayes’ rule [27]. Discriminant analysis works by creating one or more linear combinations of variables, creating a new latent variable for each function. These functions are called discriminant functions, and they are used for discriminating the classes. The model fits a Gaussian density to each class, assuming that all classes share the same covariance matrix. To use this model as a classifier, we need to estimate from the training data the class priors, the class means, and the covariance matrices. LDA classifier is attractive because it has closed-form solutions that can be easily computed, and it inherently supports multiclass classification.

#### 2.8.5. Decision Tree

Decision Tree (DT) is a classifier that uses a tree-like graph structure composing of nodes and branches [28]. The internal nodes of the tree represent a test on a feature, each branch of the tree represents a possible decision, and the end nodes (i.e., leaves) are the output classes. DT splits the nodes on all available features and then selects the split according to an impurity measure (e.g., Gini index or entropy) in an effort to group samples with the same class together. A special type of DT is the Classification and Regression Tree (CART), which is constructed by splitting a node into two-child nodes recursively, beginning with the root node that contains the whole learning sample. CART constructs binary trees using the features and a threshold that yield the largest information gain at each node. In our study, we used DT classifier based on CART algorithm. The potential problem of a DT classifier is that a child node of a tree-structured classifier loses distribution information from the entire dataset. DT classifiers are, therefore, can be unstable because small variations in the data might result in a completely different tree being generated, and effect predictive performance. This is called high variance, which can be lowered by ensemble methods with bagging and boosting such as RF, gradient boosting or adaptive boosting.

#### 2.8.6. Random Forest

Random Forest (RF) is one of the ensemble learning methods that has proven to be a powerful technique for high-dimensional classification and skewed problems [29]. Ensemble method combines multiple separate models to improve the predictive performance and reduce variance. RF works by constructing multiple DTs and yields an ensemble classification model. In this method, every DT classifier in the ensemble is trained on an independently drawn random bootstrap sample of input data. To classify a new sample, every DT classifier produces a vote for the class it predicts, and the sample is labeled as a member of the class with the most votes.

#### 2.8.7. Adaptive Boosting

Adaptive Boosting (AdaBoost) classifier is an ensemble method that fits a sequence of weak learners (i.e., models that are only slightly better than random guessing, such as small DTs) on iteratively modified versions of the training data [30]. The predictions from all of the learners are then combined through a weighted majority vote (or sum) to produce the final prediction. In each iteration, the weights associated to the observations are individually updated, and the learning algorithm is reapplied to the data. Observations that were incorrectly misclassified by the model in the previous iteration have their weights increased, whereas the weights are decreased for those that were predicted correctly. As iterations proceed, observations that are difficult to predict are given a higher probability of being selected for the next iteration in the sequence. Each subsequent weak learner is thereby forced to concentrate on the observations that are misclassified in the previous ensemble. Generalization of AdaBoost for multiclass classification has also been introduced, which is referred as AdaBoost-SAMME [31]. In our study, we used CART DT as weak learner in the AdaBoost-SAMME model.

#### 2.8.8. Gradient Boosting

Gradient Boosting (GB) classifier builds an ensemble of many weak learners (e.g., typically DTs) that are built sequentially from residual-like measures from the previous DT [32]. In each iteration, a DT is built from a random subsample of the dataset, selected without replacement. Finally, all the weak learners are added together as a weighted sum of the weak learners. The overall model accuracy gets progressively better with each additional weak learner. Similar to other boosting algorithms, GB classifier builds the additive model in a greedy fashion where the newly added DT tries to minimize the loss, given the previous ensemble model.

#### 2.8.9. Performance metrics and measurement tools

We presented our results using basic evaluation metrics derived from confusion matrix associated with the classifier. Confusion matrix is a table of correct and incorrect prediction results of a classifier model on a set of data for which the true values are known. Based on the confusion matrix, we reported the following metrics: accuracy (ACC), Sensitivity (SENS), Specificity (SPEC), and Precision (PREC). We employed F1 Score that combines precision and recall (i.e., sensitivity) as a measure of effectiveness of classification. We also employed average per class F1 score (F1_m_) for multiclass or binary classification setting, which is calculated by averaging F1 scores over all classes.

Furthermore, we reported Receiver Operating Characteristics (ROC) Curve and associated area under the curve value (ROC AUC) for the models. ROC curves were produced using F1_m_ score, cross-validation (CV) scheme, and the complete T&V dataset. CV scheme was selected to be based on stratified 5 fold split with training and validation sets that were created while preserving class proportions. Taking all of these 5 curves, it is possible to calculate the mean AUC, and see the variance of the curve when the training set is split into different subsets. This roughly shows how the classifier output is affected by changes in the training data.

We presented precision-recall curve (PRC) and associated area under the curve (PRC AUC) values for the models. PRC shows the trade-off between precision and recall for different thresholds.

We also reported learning curves to compare the performance of a model on training and validation data over a varying number of training data samples. It is a useful tool to find out how much we benefit from adding more training data, and whether the classifier suffers more from a variance error or a bias error. Learning curves for the RF models were produced using CV scheme using the complete T&V dataset. CV scheme was selected to be based on stratified random sampling with replacement in which 20 random splits with 70% training and 30% validation sets with preserved class proportions. Afterwards, the scores were averaged over all 20 runs for each training subset size and plotted against the varying data size.

In diagnosis of CS and identification of its cause, feature importance might be a useful tool by which one is able to interpret the outcome of the model and determine the most impactful medical tests. Therefore we present relative importance of features in RF models using mean decrease in impurity calculation [33].

The performance evaluation metrics and their equations are given in Table 3. Detailed description of performance evaluation metrics and tools used in this study are given in Section 1.4 of the Supplementary Materials.

**Table 3.**
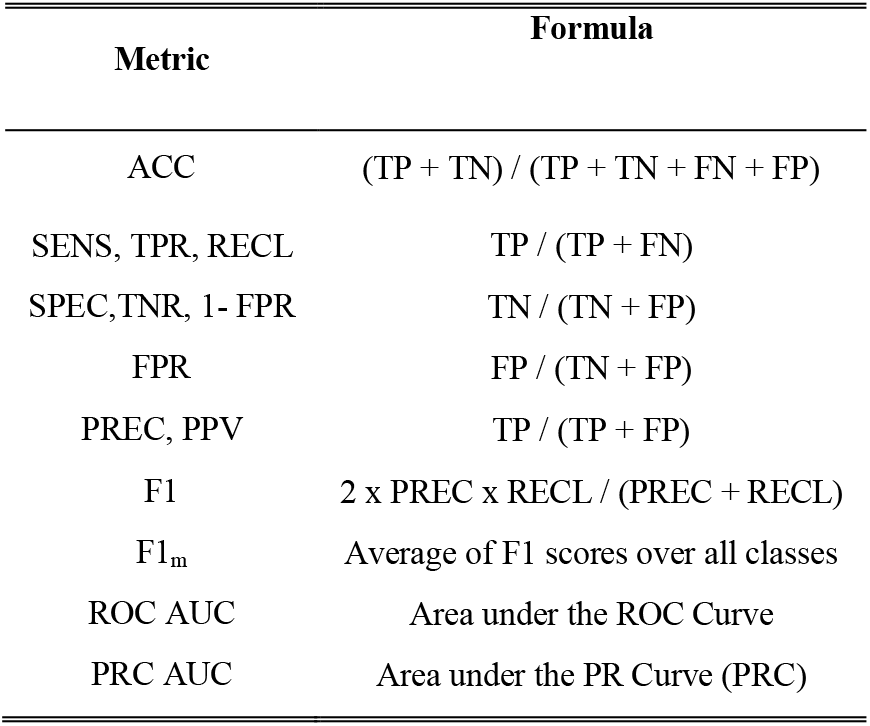
Performance evaluation metrics.

| Metric | Formula |
| --- | --- |
| ACC | $(TP + TN) / (TP + TN + FN + FP)$ |
| SENS, TPR, RECL | $TP / (TP + FN)$ |
| SPEC, TNR, 1- FPR | $TN / (TN + FP)$ |
| FPR | $FP / (TN + FP)$ |
| PREC, PPV | $TP / (TP + FP)$ |
| F1 | $2 \times PREC \times RECL / (PREC + RECL)$ |
| $F1_m$ | Average of F1 scores over all classes |
| ROC AUC | Area under the ROC Curve |
| PRC AUC | Area under the PR Curve (PRC) |

### 2.9. Definition, comparison, and selection of algorithms

In this study, we aim to evaluate different algorithms in terms of their predictive performance (i.e., generalization accuracy) on unseen independent data and identify the ML algorithm that is best suited for the diagnosis of CS. We compared trained models from the algorithm’s model space and selected the best performing models by tweaking the hyperparameters of the algorithm. Furthermore, we aim to optimize hyperparameters from a given hyperparameter space by comparing and selecting values of hyperparameters with respect to their performance in minimizing error. NCV approach is well suitable for these tasks under limited data size and produces almost unbiased performance estimates [34]. NCV is relatively straightforward as it is a nesting of two k-fold CV loops: the inner loop is responsible for the model selection and parameter optimization, and the outer loop is responsible for estimation of the generalization accuracy for model evaluation.

We created classifiers for these class comparison strategies (n = 11): (i) multiclass comparison of all types (i.e., NF, SC, AD, and PT), (ii) one vs. one binary comparisons (PTvsAD, ADvsNF, PTvsNF, SCvsNF, ADvsSC, PTvsSC), and (iii) one vs. all binary comparisons (SCvsALL, PTvsALL, ADvsALL, and ALLvsNF). Please note that ALL refers to all classes in multiclass setting whereas it refers to the rest of the classes in binary setting. This means that class labels were merged together for classes in the same comparison. Class comparisons were iterated for the features of diagnostic Stage-1 and diagnostic Stage-2. In the end, 22 classifier models were evaluated per each algorithm.

We evaluated each algorithm and its associated models with a 5×3 NCV procedure with steps as the following: the input T&V dataset was divided into stratified 5 folds (i.e., each fold contains approximately the same percentage of samples of each target class as in the input dataset.). For each iteration in the outer loop, 1 data fold was reserved as held-out validation data for model evaluation. The remaining 4 folds were passed to the inner loop where model parameter tuning was performed. All the models have hyperparameters that must be compared and selected. These include, for example, penalty term for overfitting in Logistic Regression, the kernel coefficient in SVM, and the number and depth of trees in RF. The parameter values evaluated for each model are given in Table S3 in the Supplementary Materials. The input data passed from the outer fold was divided into stratified 3 folds. The inner loop included a grid search over candidate hyperparameters using 2 folds of data. Each parameter setting was evaluated with the remaining 1 fold validation data. Inner loop was repeated 3 times, each time holding out a different validation fold. The hyperparameters that yielded the best average CV score were selected and reported back to the outer loop. The model was then trained on the data in the 4 folds using the best parameters passed from the inner loop and then evaluated for its predictive performance using the 1 fold held-out validation data in the outer loop. This process was repeated 5 times in the outer loop, resulting in evaluations of model performance for 5 times, and the average evaluation score was obtained from averaging outer held-out validation datasets. Scores using several evaluation metrics were reported. NCV procedure was repeated for each of the classification algorithms (n = 8) and for each class comparison strategy (n = 11) using a different feature set for each of the diagnostic stages (n = 2). Overall, NCV was performed 176 times. The best performing algorithm was selected according to the highest grand average for all runs.

### 2.10. Training, testing, and evaluation

Following the algorithm selection, the selected algorithm (i.e., RF in our study) was used to build classification models with different class comparison strategies and feature sets which were selected after discussing it with physicians specialized in CS as to how useful it will be in clinical diagnosis and prognosis. We created classifiers for multiclass comparison for all class types (NF, SC, AD, and PT), one vs. one binary comparisons: PTvsAD, ADvsNF, PTvsNF, SCvsNF, ADvsSC, PTvsSC, and one vs. all binary comparisons: SCvsALL, PTvsALL, ADvsALL, and ALLvsNF. ALL refers to that class labels are merged together for classes in the same group of comparison. Class comparisons are iterated for the features of diagnostic Stage-1 and diagnostic Stage-2.

The hyperparameters used in the models were determined by 3-fold CV hyperparameter search approach similar to the one in algorithm selection procedure. However, this time, all of the T&V dataset was used in the search.

In the training phase, ML model was inferred from T&V dataset with known class labels. The parameters of the model were optimized by fitting the observations in the dataset to the output target variable. Afterward, the trained models were evaluated on the FIT dataset for their predictive performance (i.e., generalization accuracy) for unseen data. At the final step, complete dataset was used to train final classification models to be used for testing new samples.

## 3. Results

### 3.1. Algorithm Comparison

Table 4 summarizes the algorithms comparison results in terms of performance estimates in classification of each of 4 classes (i.e., NF, SC, AD, and PT) computed by 5×3 NCV procedure on the T&V dataset. It includes average per class F1 scores (i.e., average of the F1 scores calculated for each class of the target variable) and accuracy scores of the Stage-1 and Stage-2 models, and overall average scores over all models (n = 22) in both Stage-1 and Stage-2. Each score in Stage-1 and Stage-2 columns in Table 4 is the mean of performance scores averaged over validation folds in 5×3 NCV for each model (n = 11) created for all class comparison strategies.

**Table 4.** Overall results of algorithm comparison.

| Algorithm | Stage-1 Scores<br>(5 features) |  | Stage-2 Scores<br>(9 features) |  | Overall Average Scores |  |
| --- | --- | --- | --- | --- | --- | --- |
|  | F1<sub>m</sub> | ACC | F1<sub>m</sub> | ACC | F1<sub>m</sub> | ACC |
| Random Forest | 88.1±9 | 90.3±8 | 90.9±8 | 92.5±7 | 89.5±9 | 91.4±8 |
| SVC | 85.5±11 | 87.7±10 | 88.7±9 | 90.6±7 | 87.1±10 | 89.2±9 |
| Logistic Regression | 85.4±13 | 87.5±11 | 88.2±10 | 90.2±8 | 86.8±11 | 88.8±10 |
| AdaBoost | 85.5±10 | 87.9±9 | 87.3±11 | 89.4±10 | 86.4±11 | 88.6±10 |
| Linear Discriminant Analysis | 83.7±15 | 87.9±9 | 88.5±12 | 90.8±9 | 86.1±14 | 89.4±9 |
| K-nearest Neighbor | 84.3±12 | 87.2±10 | 87.7±11 | 90.2±9 | 86±11 | 88.7±10 |
| Decision Tree | 84.7±11 | 87.2±10 | 87.2±11 | 89.1±10 | 86±11 | 88.2±10 |
| Gradient Boosting | 82.2±11 | 85.6±10 | 85±12 | 87.6±11 | 83.6±12 | 86.6±10 |

The highest achieved F1_m_ score for RF was 90.9±8 with Stage-2 setting of 9 features whereas it was 88.1±9 with Stage-1 setting of 5 features. RF had an overall average score of 89.5±9. Other algorithms had comparable average scores to RF algorithm. However, RF models had smaller standard deviations. The highest deviations were seen in Linear Discriminant Analysis. It could be seen from these results that models built using RF showed better performance compared to the other algorithms for both Stage-1 and Stage-2 diagnosis of CS. It was also observed that model performance increases with the increase of number of features. This is an indication that important features added to the model building promote the predictive power. Also, it could be seen that F1_m_ score was generally lower than the accuracy metric. F1_m_ was used as the primary metric for comparisons throughout the study.

Table 5 gives the decomposition of NCV scores of ML algorithms averaged over Stage-1 and Stage-2 against class comparison strategies. In 6 of 11 class comparison strategies, RF ranked first. The highest score for RF was 97.8±7 achieved in PTvsNF comparison whereas the lowest score was 81.3±4 for multiclass comparison ALL. Linear Discriminant Analysis achieved the highest score in PTvsAD and PTvsALL comparisons. Logistic Regression ranked first in PTvsSC, ADvsNF, and in multiclass ALL comparison. DT ranked first in SCvsNF comparison. It was also seen that SCvsALL, ADvsSC, and multiclass ALL classifications were the cases in which algorithms generally achieved lower performance.

**Table 5.** Comparison of algorithms against class comparison strategy.

|  | ALL | ALL vsNF | SCvs ALL | PTvs ALL | ADvs ALL | PTvs AD | ADvs NF | PTvs NF | SCvs NF | ADvs SC | PTvs SC |
| --- | --- | --- | --- | --- | --- | --- | --- | --- | --- | --- | --- |
| Algorithm | $F1_m$ % | | | | | | | | | | |
| Random Forest | **81.3±4** | **91.7±4** | **81.4±8** | 93.3±8 | **86.4±7** | 93.1±12 | 93.2±7 | **97.8±7** | 90.8±8 | **82.1±9** | 93.5±5 |
| SVC | 78.3±5 | 89.6±5 | 74.6±11 | 92.1±6 | 83±8 | 95±10 | 92.6±7 | 95.1±7 | 85.5±7 | 80.8±8 | 91.9±8 |
| Logistic Regression | **81.3±4** | 88.1±4 | 67.4±9 | 94.4±6 | 79.6±9 | 95.9±10 | **94.1±6** | 96.7±6 | 85.1±9 | 78.5±8 | **93.6±8** |
| AdaBoost | 70.9±8 | 90.5±3 | 80±6 | 91.5±8 | 85.4±6 | 90±14 | 92.7±7 | 93.7±7 | 90.3±5 | 80.5±9 | 84.6±15 |
| Linear Discriminant Analysis | 80.6±5 | 89.5±4 | 59±14 | **96.1±5** | 77.6±14 | **97.1±7** | 93.4±6 | 97.2±7 | 84.8±9 | 78.9±10 | 92.6±7 |
| K-nearest Neighbor | 75.8±5 | 89.6±5 | 73.4±9 | 92.7±9 | 78.6±8 | 96.2±7 | 91.3±8 | 97.2±7 | 81.4±9 | 76.3±11 | 93.5±8 |
| Decision Tree | 75.9±9 | 86.6±6 | 74±10 | 89.7±9 | 84.1±7 | 90.1±13 | 90.3±9 | 94.3±8 | **91.9±7** | 77.8±15 | 91.1±4 |
| Gradient Boosting | 74.6±6 | 88.2±3 | 71.1±11 | 89.1±9 | 78.6±10 | 89.2±12 | 88.9±10 | 94.2±8 | 83.6±8 | 71.8±15 | 90.4±5 |

According to NCV results, RF algorithm was found to be the best performing algorithm and selected for in-depth study. A subset of RF models from 11 different class comparison strategies was selected for training after discussing the clinical methods and models with physicians specialized in CS. As aforementioned in Section 1.1 (see also detailed information in Section 1.2 of Supplementary Materials), usually at Stage-1, it is aimed to diagnose patients with hypercortisolism. Therefore, discrimination between nonfunctional adrenal adenoma and CS regardless of its subtype is required. Therefore, ALLvsNF one vs. all binary classification model with Stage-1 features was employed. At the second stage, identification of specific CS subtype against the rest of alternative CS subtypes is required. Therefore, SCvsALL, ADvsALL, and PTvsALL one vs. all binary models with Stage-2 features were employed. Multiclass classification was employed with both Stage-1 and Stage-2 features.

### 3.2. Parameter Optimization

Table 6 shows the optimized hyperparameters and mean performance scores obtained from 3-fold cross-validated exhaustive search over hyperparameter values for selected RF models using the complete T&V dataset. Optimized hyperparameters were the number of decision trees employed in the RF, max depth of decision trees, split criterion, and class weight criterion.

**Table 6.**
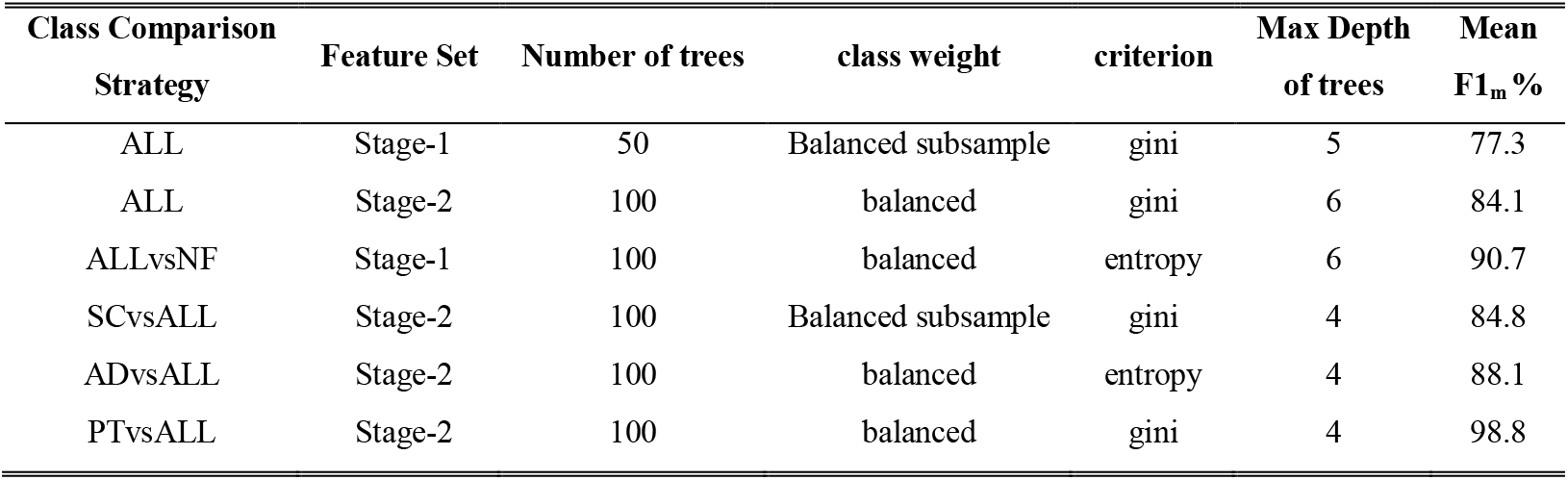
Best hyperparameters for RF models obtained from cross validated grid search.

| Class Comparison Strategy | Feature Set | Number of trees | class weight | criterion | Max Depth of trees | Mean F1<sub>m</sub> % |
| --- | --- | --- | --- | --- | --- | --- |
| ALL | Stage-1 | 50 | Balanced subsample | gini | 5 | 77.3 |
| ALL | Stage-2 | 100 | balanced | gini | 6 | 84.1 |
| ALLvsNF | Stage-1 | 100 | balanced | entropy | 6 | 90.7 |
| SCvsALL | Stage-2 | 100 | Balanced subsample | gini | 4 | 84.8 |
| ADvsALL | Stage-2 | 100 | balanced | entropy | 4 | 88.1 |
| PTvsALL | Stage-2 | 100 | balanced | gini | 4 | 98.8 |

### 3.3. Training

Table 7 shows the performance results of the trained RF models. PTvsALL achieved the highest classification score (F1_m_) of 100% whereas ADvsALL had the lowest score of 90.3%. High sensitivity and specificity were achieved in all models, ranging from 92.7% to 100% and from 93.7% to 100%, respectively. ALLvsNF model had 100% sensitivity, and was able to correctly catch all CS samples at Stage-1. It also correctly generated a negative result for NF samples at 95.9% specificity. Precision or Positive Predictive Power (PPV) of ALLvsNF model was 96.9% that means only 3.1 % of positive results generated by the model was actually NF. Multiclass ALL model at Stage-1 had 95% classification score (F1_m_) and achieved sensitivity values above 92%, specificity values above 97%, precision values above 87%, depending on the class type. AD type had the lowest precision of 87.5% compared to other types in the multiclass ALL model. Multiclass ALL model at Stage-2 achieved 96.4% classification score (F1_m_) and improved on sensitivity and specificity compared to the multiclass ALL model at Stage-1. Confusion matrix in Fig. 2(a) shows the number of correct predictions as well as false positives and false negatives for Stage-2 multiclass classification models in training phase. SCvsALL model had sensitivity of 97.6%, specificity of 93.7%, and precision of 83.3%. ADvsALL model had sensitivity of 100%, specificity of 95%, and precision of 80.6%. The model was able to recognize AD samples. PTvsALL model had sensitivity of 100%, specificity of 100%, and precision of 100%. The model was able to correctly discriminate PT type and the rest of the classes.

**Table 7.**
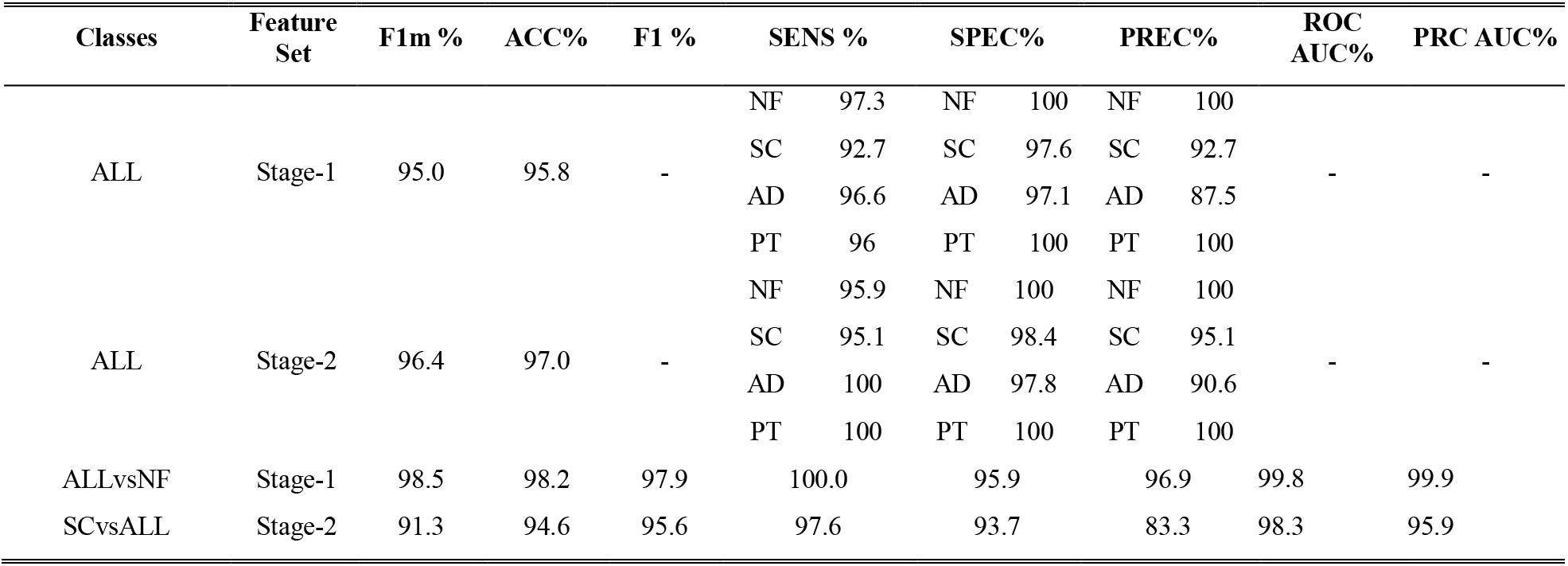
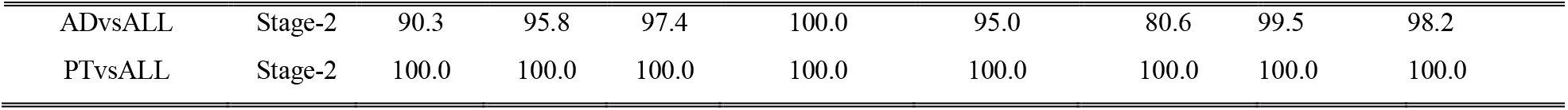
Training results of RF models.

**Fig. 2.**
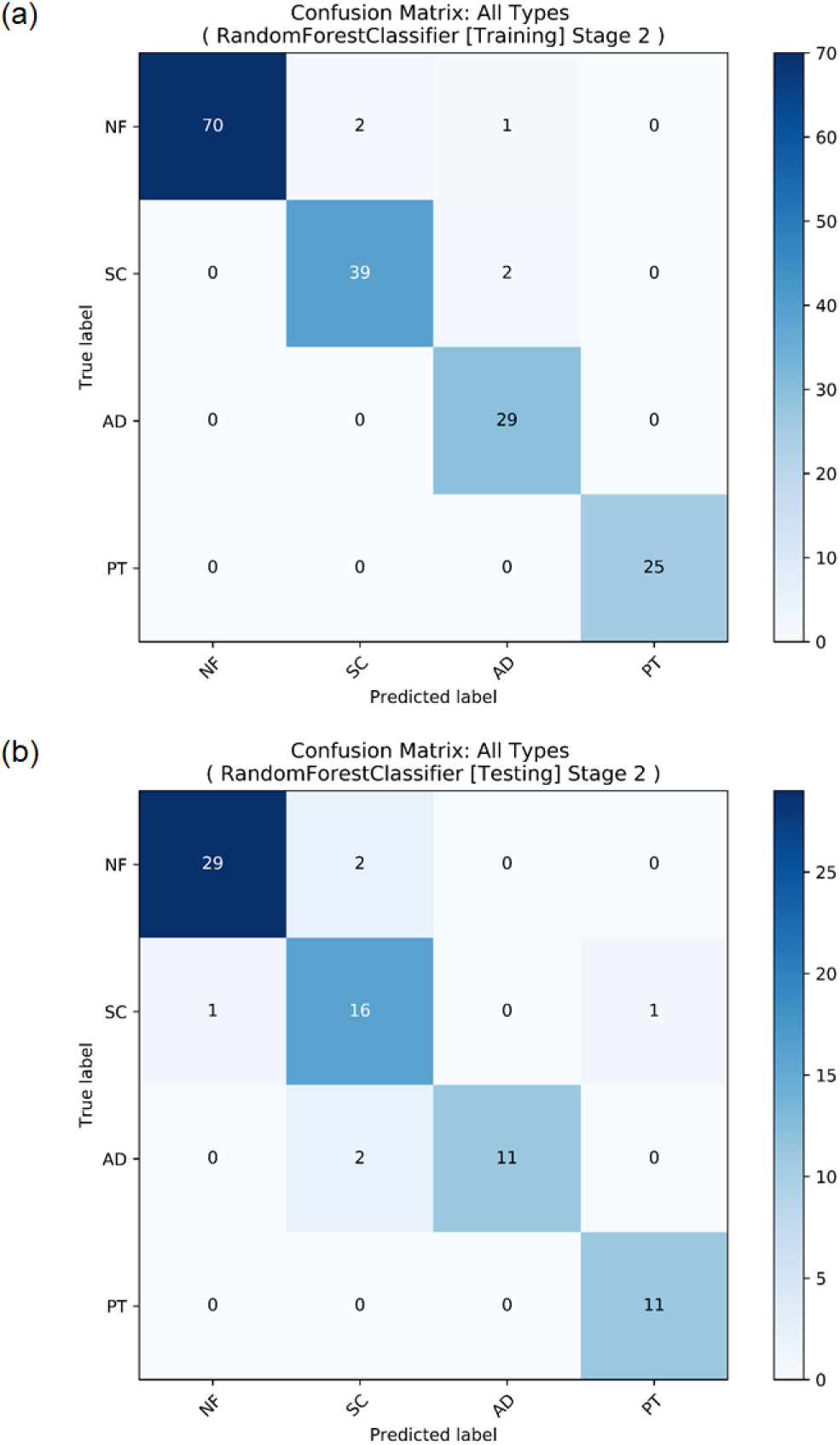
Confusion matrices of RF multiclass ALL model at Stage-2 in (a) training phase and (b) testing phase.

Fig. 3 shows cross-validated learning curves of ALLvsNF model at Stage-1 and Multiclass ALL model at Stage-2 in training phase. General characteristics of the curves look promising. For multiclass ALL model at Stage-2, the curve shows good performance. However, there is a significant variance between the training and validation curves of the model. High score in the training curves starting from the low data sizes indicates low bias, and this is usually typical for tree-based algorithms. Training and validation curves appear to be converging but there is significant variance (i.e., the gap between curves) in both models. These characteristics show overfitting in the learning procedure, and the rate of convergence is observed to be relatively small. Similar characteristics are also seen in learning curves of ALLvsNF model, but the variance is smaller compared to learning curve of the multiclass ALL model. For SCvsALL, ADvsALL, and PTvsALL models at Stage-1, the rate of convergence is higher compared to the multiclass model and ALLvsNF models, and multiclass ALL model at Stage-1 show similar characteristics to its Stage-2 counterpart, as seen in Fig. S2 of Supplementary Materials. In conclusion, learning curves show very good performance scores with some variance. Adding more informative samples to the dataset would probably help to improve the model fitting and increase generalization accuracy, given the current settings in the learning procedure.

**Fig. 3.**
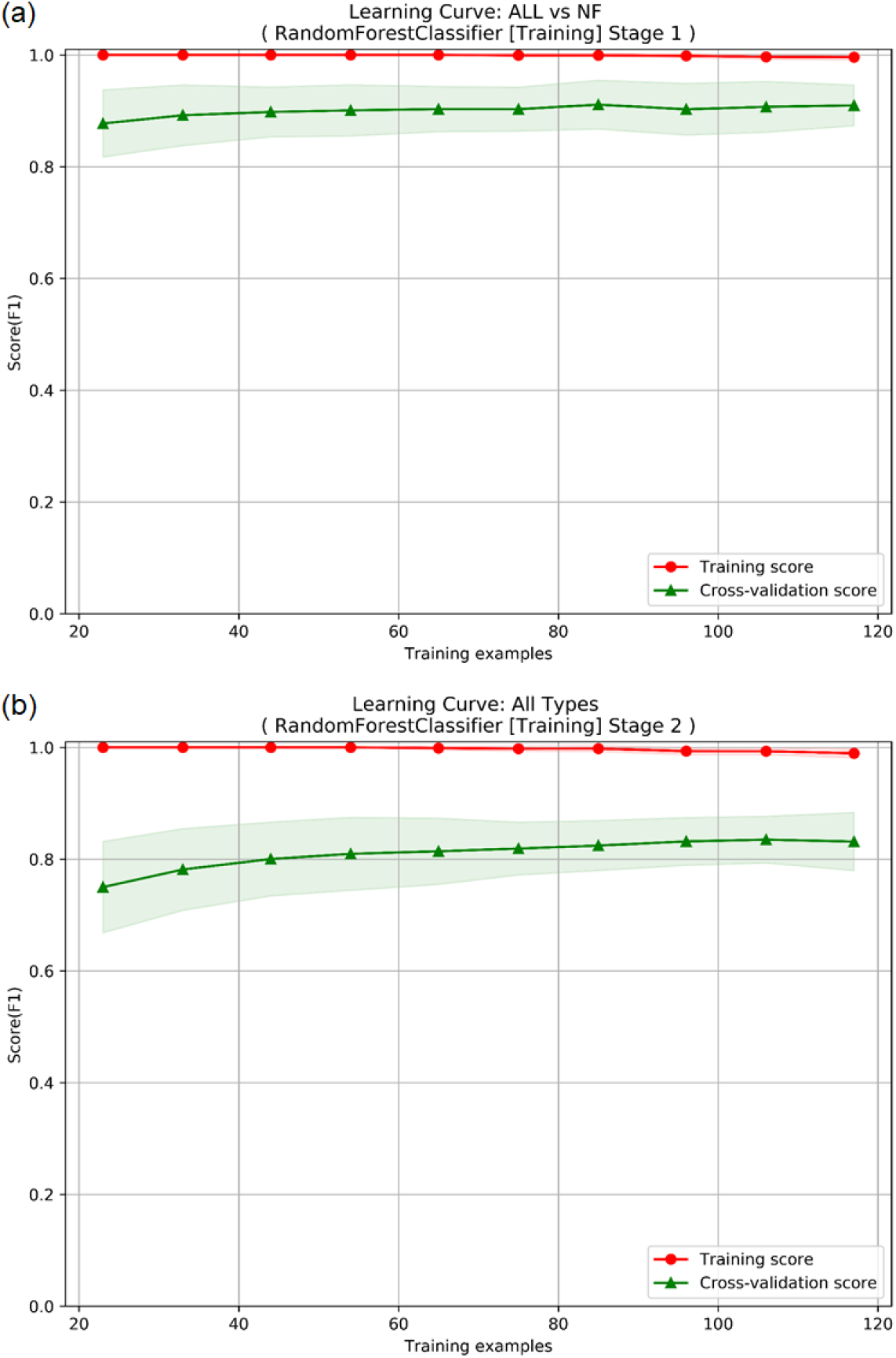
Learning curves of (a) RF ALL vs. NF model at Stage-1 and (b) multiclass ALL model at Stage-2.

The cross-validated ROC Curve and Precision-Recall (PR) Curve are shown in Fig. 4 for ALLvsNF model at Stage-1. For SCvsALL, ADvsALL, and PTvsALL models at Stage-2, the ROC curves are given in Fig. S3 of Supplementary Materials. ROC curves use different subsets of the T&V dataset, created from 5-fold cross-validation. Accuracy is measured by the area under the ROC curve (AUC). All curves have high mean AUC values: 0.9690±0.017 for ALLvsNF, 0.947±0.074 for ADvsALL, 0.926±0.022 for SCvsALL, and 0.995±0.000 for PTvsALL. This means that classifiers are better at classifying positive and negative observations. CV roughly shows how the classifier output is affected by changes in the training data. Standard deviations calculated for CV folds indicate that PTvsALL model is more robust to changes in the data whereas ADvsALL model is more susceptible to data perturbations.

**Fig. 4.**
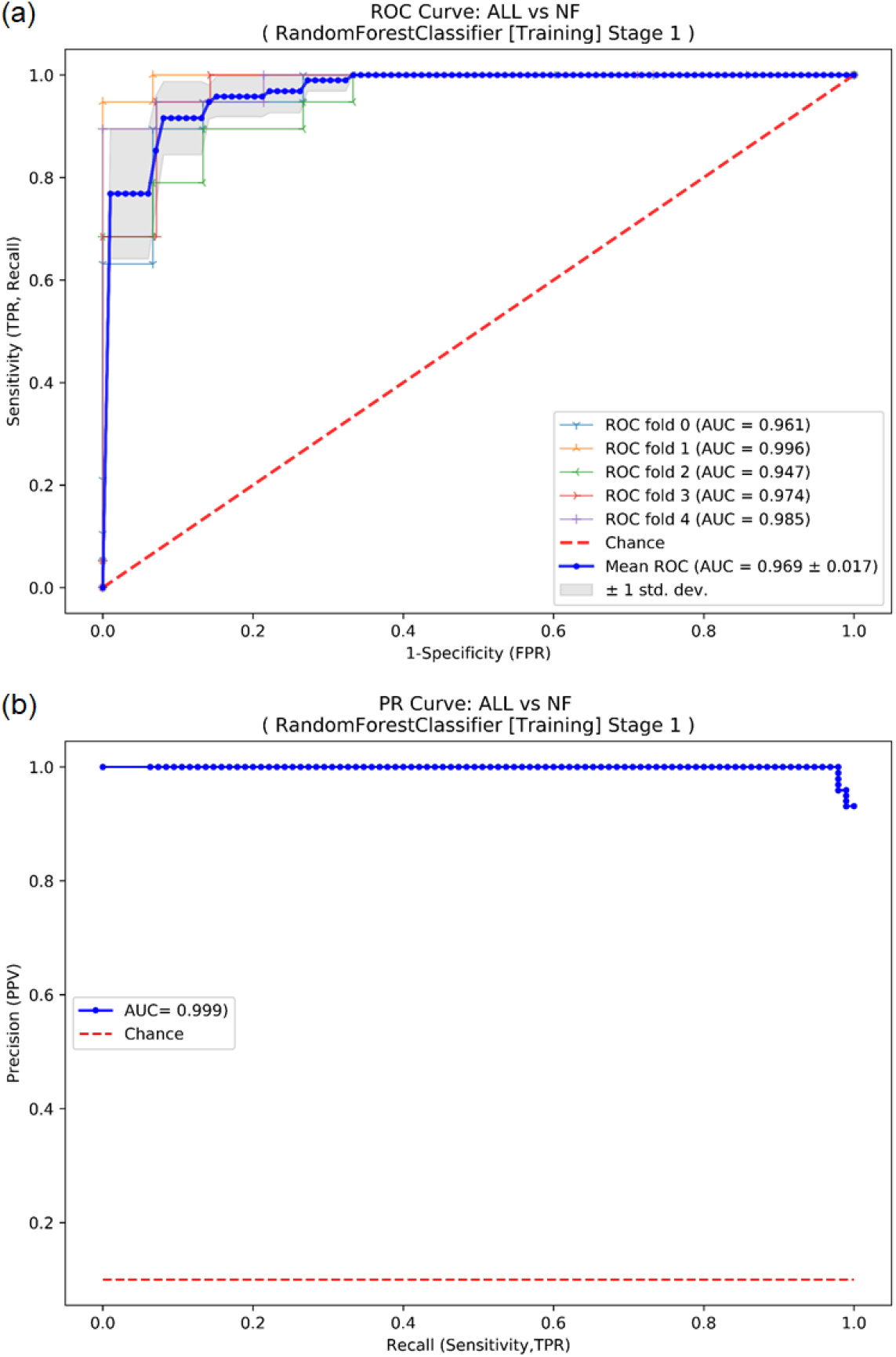
(a) Cross-validated ROC Curve and (b) PR Curve of RF ALL vs. NF binary model at Stage-1 in training phase.

### 3.4. Feature Importance

Fig. 5 shows relative feature importance levels of trained models. For multiclass ALL model at Stage-1, the features 1mgDSTc, bacht, and mc were found to be the relatively most important features that contribute most to the classification performance. For ALLvsNF model, 1mgDSTc was by far the most important feature and mc being the second. Relatively most important features for multiclass ALL model at Stage-2 were inferred to be 1mgDSTc, bacth, pitMass, and mc. For SCvsALL model, 1mgDSTc, 2mgDSTc, bacth and mc rank at top. The features mc, 1mgDSTc, and bacth were found to be the most important features for ADvsALL model, For PTvsALL model, pitMass was by far the most important feature, accompanied by bacth and 8mgDSTc.

**Fig. 5.**
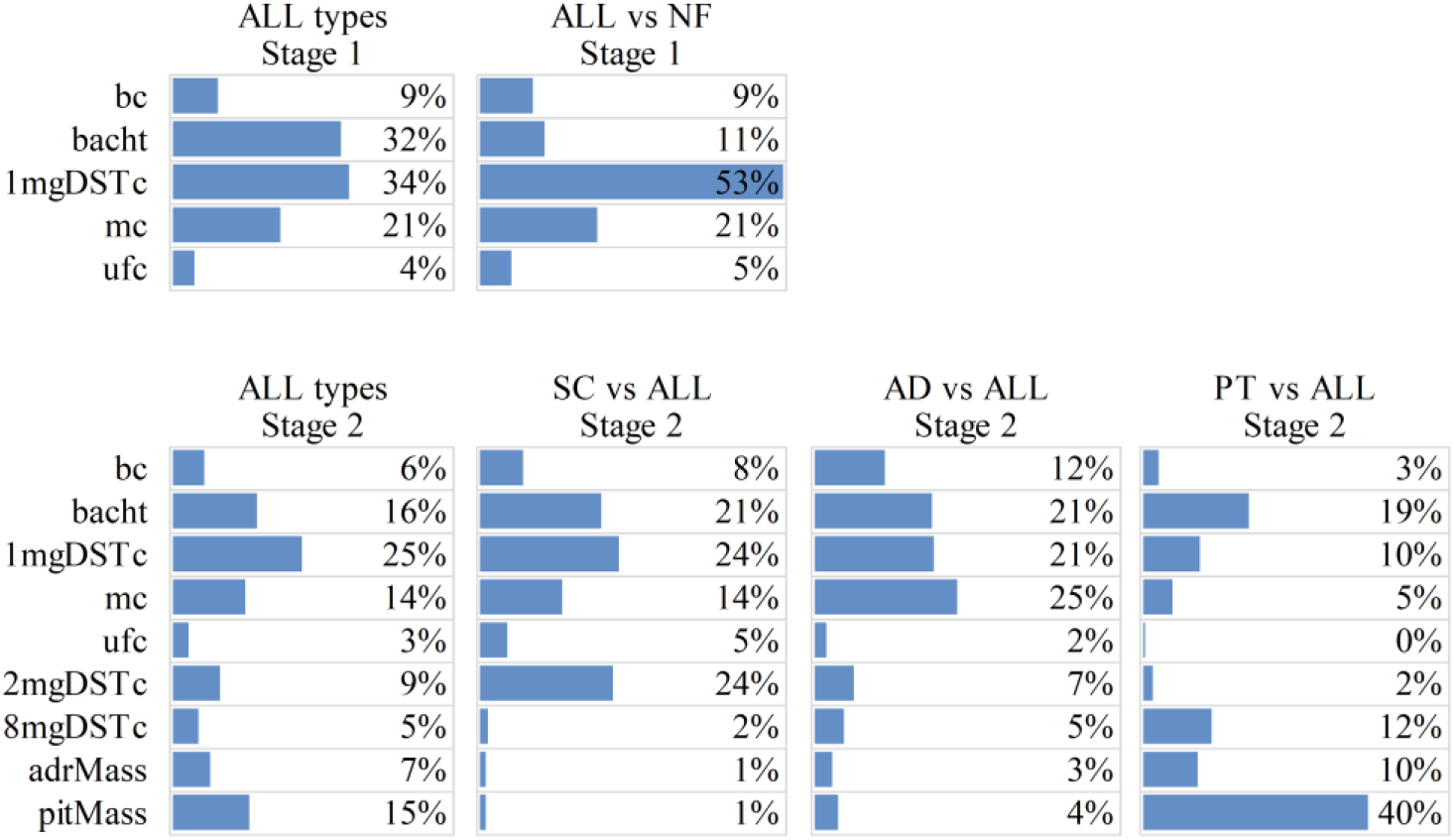
Feature importance levels of RF models.

### 3.5. Final Testing

Table 8 shows the results of the testing of trained models using FIT dataset to show their generalization accuracy to unseen independent samples.

**Table 8.**
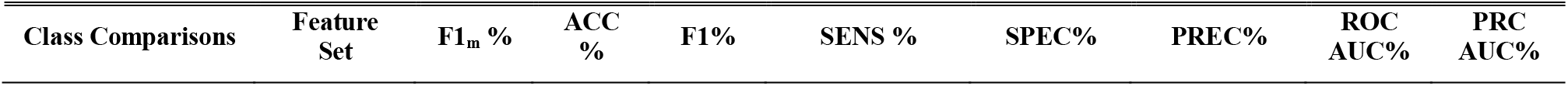
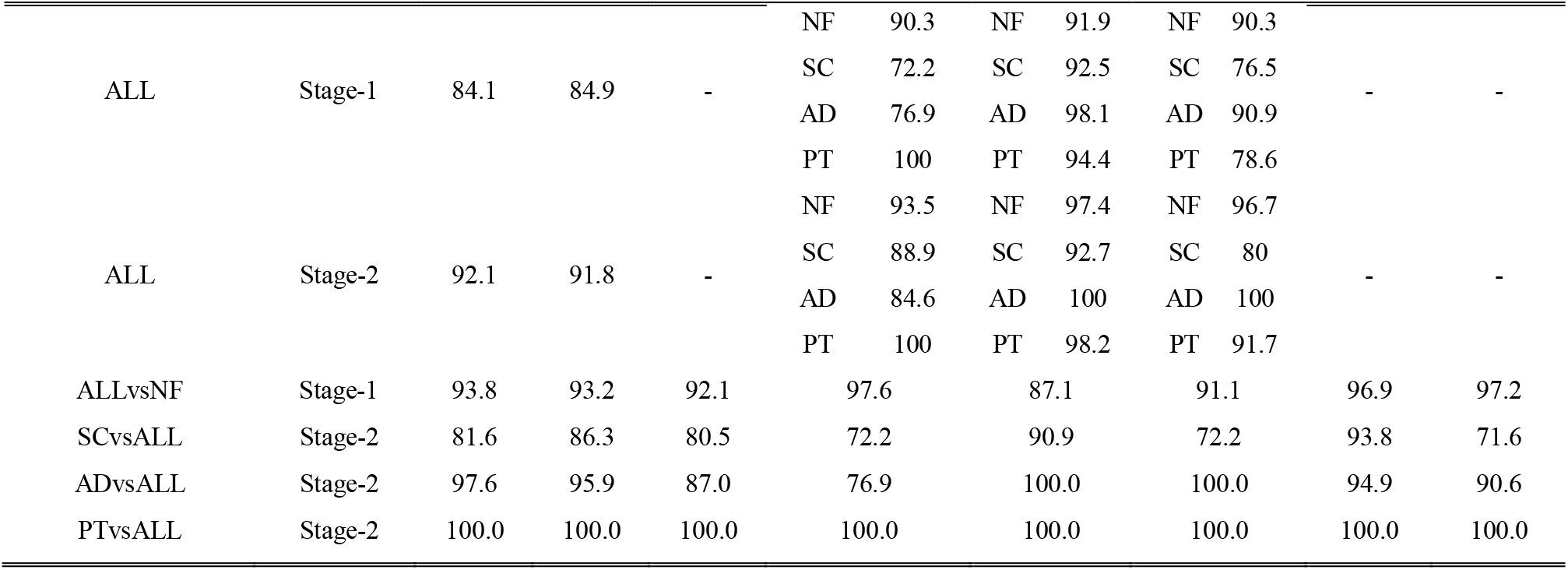
Test results of RF models.

ALLvsNF model at Stage-1 achieved 93.8% classification performance (F1_m_), and was able to correctly catch CS samples with sensitivity of 97.6%. It also correctly generated a negative result for NF samples at 87.1% specificity. Precision of ALLvsNF model was 91.1% which means 8.9% of positive results generated by the model were actually NF. Fig. 6 shows ROC curve and PR curve of ALL vs. NF binary model at Stage-1 in test phase. ROC curve has AUC of 0.969 which means there is 96.9% chance that model is able to distinguish between classes. PR curve has AUC of 0.972 which represents both high recall and high precision.

**Fig. 6.**
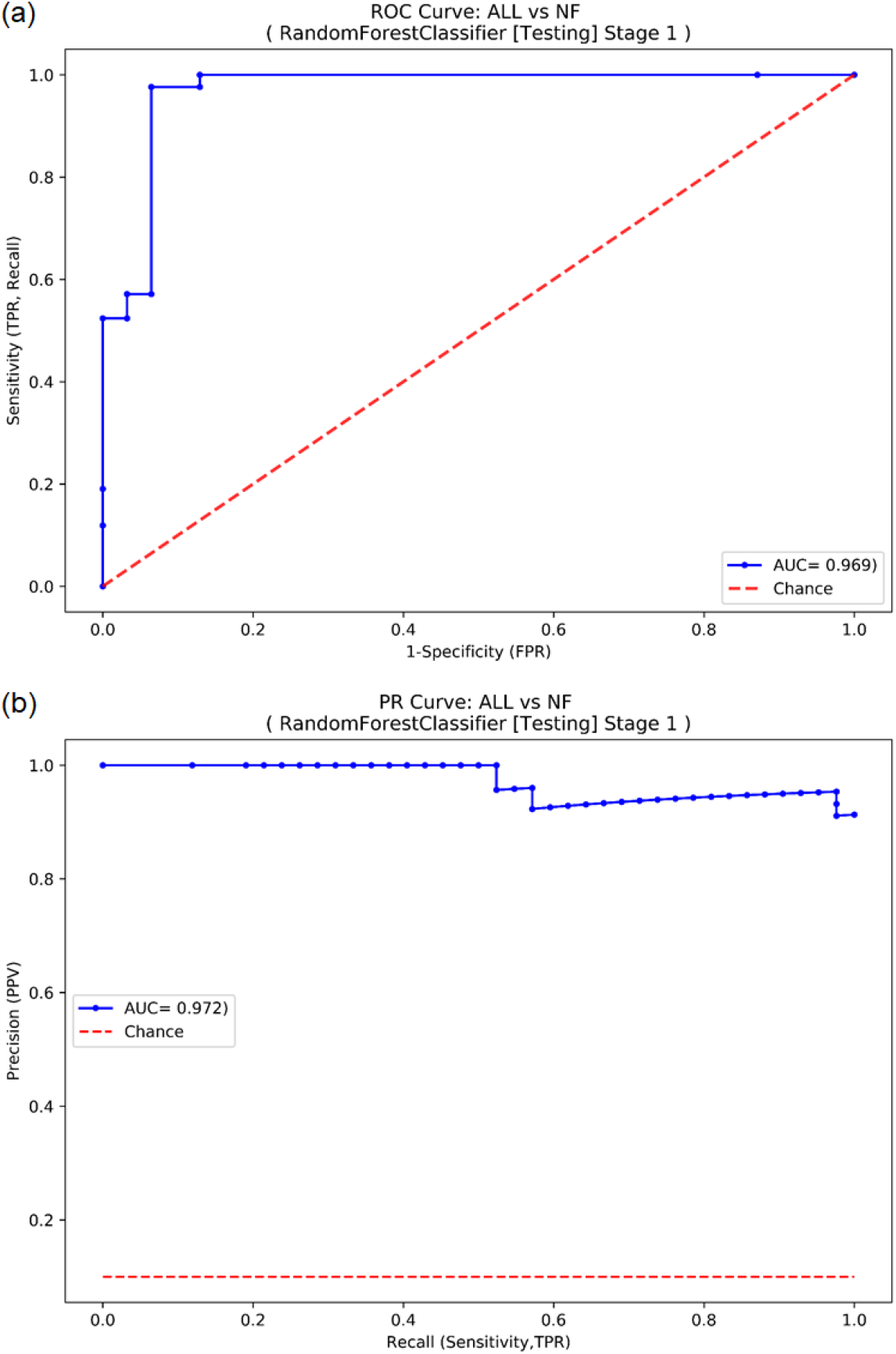
(a) ROC Curve and (b) PR Curve of RF ALL vs. NF binary model at Stage-1 in test phase.

Multiclass ALL model at Stage-1 had 84.1% classification score (F1_m_). The sensitivity value for PT class was 100%, which means that multiclass ALL model was able to catch all PT samples. Sensitivity for NF was 90.3%. However, the multiclass ALL model at Stage-1 had moderate sensitivity values of 72.2% and 76.9% for SC and AD, respectively. All specificity values for classes were high, ranging from 91.9% to 98.1%. Precision values for NF and AD were above 90%. However, precision values were moderate for SC and PT. Multiclass ALL model at Stage-2 improved in all aspects compared to the Stage-1 multiclass ALL model. It achieved 92.1% classification score (F1_m_) and a range from 84.6% to 100% per class. Average per class sensitivity (i.e., average of sensitivity over classes) was 91.8%. Specificity ranged from 92.7% to 100% per class, and average per class specificity was 97.1%. Precision was between 80% and 100% per class, and average per class precision was 92.1%. Fig. 2(b) shows the confusion matrix of multiclass classification model at Stage-2 in testing phase. It indicates the number of correct predictions as well as false positives and false negatives.

SCvsALL model had 81.6% classification performance score (F1_m_), a moderate sensitivity of 72.2%, high specificity of 90.9%, and moderate precision of 72.2%. This moderate performance can also be observed in PR curves given in Fig. S5 of Supplementary Materials. It is also seen ADvsALL model had high classification performance (F1_m_) of 97.6%, moderate sensitivity of 76.9%, maximum specificity of 100%, and maximum precision of 100%. PTvsALL model had maximum score, sensitivity, specificity and precision of 100%. The model was able to correctly discriminate all PT type samples and samples of the rest of the classes.

### 3.6. Clinical Performance Evaluation

Clinical performance of ML models was evaluated by comparing their predictions to the judgments made by human experts as shown in Table 9. For this comparison, patient data of 73 cases (i.e. FIT dataset) which were not used in model development were divided randomly into three subsets. Three physicians specialized in CS were asked to make Stage-1 and Stage-2 diagnoses using the patients’ test results in each subset. Experts tried to separate CS from nonfunctional adrenal adenoma in the biochemical screening testing stage (Stage-1). They were able to classify subjects with F1 score of 76.6 % whereas F1 score of ALLvsNF model was 98.5%. In follow-up testing setting (Stage-2), experts were asked to discriminate among subtypes of CS using biochemical tests and imaging findings of patients who are suspected of CS. The overall achievement of the experts resulted in an F1 score of 49.4% while multiclass ALL model achieved an F1 score of 96.4%. The poor score of human experts was mainly because of the failure of discriminating the cases with subclinical CS. It was found that with the lack of other information such as findings of physical anomalies, physicians had tendency to avoid labeling subclinical CS cases. When subclinical CS cases were removed, the overall performance of the human experts achieved an F1 score of 85.45% which is still lower than the performance achieved by the ML model. This revealed that ML model is able to find underlying patterns in the CS data with a less set of features (i.e. biochemical tests, presence of other illnesses or clinical findings).

**Table 9.** Clinical performance comparison of ML models vs. Human Expert.

| Predictor | Classes | Feature Set | F1m % | ACC% |
| --- | --- | --- | --- | --- |
| ML | ALL | Stage-2 | 96.4 | 97.0 |
| Human Expert | ALL | Stage-2 | 49.4 | 64.3 |
| ML | ALLvsNF | Stage-1 | 98.5 | 98.2 |
| Human Expert | ALLvsNF | Stage-1 | 76.6 | 76.7 |

## 4. Discussions

Results of ML algorithm comparison as given in Table 5 and Table 6 indicate that most of the compared algorithms achieve good estimates of predictive performances on the T&V dataset using NCV procedure. However, RF algorithm is found to be the best algorithm in overall performance according to the scores achieved for different class comparison schemes and feature sets. The algorithms generally show better performance with features of Stage-2 than with features of Stage-1. This is probably due to the inclusion of more informative features into the models, and it helps better fitting of model parameters to the data. This is actually observable in the relative feature importance levels of RF-based multiclass models that were inferred in the training phase as seen in Fig. 5. For the RF-based multiclass model at Stage-1, bacth, 1mgDSTc, and mc contributed much to the predictive power so that the model achieved classification accuracy of 95% (See Table 7). Similarly, for the RF multiclass model at Stage-1, the same features were found to be the most important features besides newly added pitMass feature, and the model achieved an improved classification accuracy of 96.4%.

Although models based on one vs. one class comparison strategy technically show good performance results at algorithm selection step, one vs. one binary models were excluded from further training and testing because of their limited clinical use. For example, for ADvsSC binary model at Stage-2, one has to eliminate other classes beforehand, and a dataset consisting of only these two classes is required for the classification results to have real meaning. Similarly, ADvsSC model at Stage-1 requires samples that contain only AD and SC types for training before making predictions using the model. This looks infeasible because one has to first eliminate NF and PT classes from the samples, which are not yet known. Moreover, the biochemical tests performed at Stage-1 are generally aimed to discriminate nonfunctional adrenal adenoma and CS regardless of its subtype. Clinical testing at Stage-2 aims to discriminate the cause of CS such as adrenal CS, ectopic CS or pituitary CS.

There are important limitations of the proposed system at its current state. Most notably, ectopic CS samples were not utilized in model training due to the lack of enough samples. Therefore, our system cannot directly classify ectopic CS. Approximately 20% of ACTH-dependent cases are ectopic CS whereas approximately 80% of ACTH-dependent CS is pituitary CS (i.e., Cushing Disease) [5], [7]. Since both pituitary CS and ectopic CS are ACTH-dependent, some of the samples predicted as pituitary CS in the original dataset may actually be ectopic CS. Therefore, further study needs to be done by the physicians to diagnose correctly. Bilateral inferior petrosal sinus sampling (BIPSS) test, imaging findings, bacth levels, and rate of suppression in cortisol level after 8mg DST are usually informative in separating ectopic CS from pituitary CS. Note that as we gather more data samples for ectopic CS, the models can be easily updated to discriminate ectopic CS as well.

The learning curves as seen in Fig. 3 indicate the presence of variance on all models. This is probably due to the limited size of the original dataset. It is likely that additional informative samples may reduce model variance and improve predictive performance. Although we continue to collect data as part of our continuing research, it is noted that accumulation of new samples needs several years as the original retrospective dataset were created in 11 years since CS is a rare disease. Bias seen in the learning curves of T&V dataset indicates that our models may be improved by adding model complexity either in the form of additional features or by replacing with more complex model architectures. Additionally, dropping relatively unimportant features from the models might improve predictive performance. As part of our future research, we will continue to collect new samples and examine the use of additional biochemical test measurements such as ACTH level after DST as a prospective approach to model improvement and perform further experiments about clinical usefulness of the approach.

The original dataset has missing values. Use of median imputation to address missing data induces bias in the relationship between features and the target variable, and may not perform as well as more elaborate methods such as KNN and multiple imputations. However, such approaches introduce additional parameters, and this may lead to errors due to unsuccessful tuning of the parameters, and eventually reduced generalizability in case of limited data size. However, as we continue to collect more data, we intend to evaluate alternative imputation methods.

Class imbalance in the dataset is known to reduce the predictive performance of a model [17]. Several methods exist to alleviate this problem, such as downsampling and oversampling methods. Since dataset size is relatively small, downsampling the majority class causes information loss. Similarly, oversampling is likely to introduce bias to the accuracy since the new data samples are generated from a few old samples, and they cannot introduce much variance to the dataset. Therefore, we address this problem by using ML algorithms that can inherently handle imbalanced data classification with class weighting in learning process.

The interpretation of medical tests to diagnose CS and classify its subtypes requires a substantial amount of resources with respect to both time and training, and is limited by the physician’s capacity to integrate numerous and complex information. Results from studies in other hospitals and medical centers may vary, and be related to factors such as laboratory errors, patient induced errors, differences between groups, age, and gender. We demonstrated that an ML-based decision support system might help. The application of an ML classifier to diagnose CS and classify its subtypes has not previously been demonstrated. However, the use of ML approach has been applied for similar purposes such as the automated interpretation of urine steroid profiles to classify normal and abnormal profiles of several metabolic conditions including CS [35], and classification of CS using gene expression data of tumor tissues [36].

There are important advantages of our proposed approach. The approach is adaptable to new data and will improve as new samples are gathered. The prediction models require very low computational resource once trained. Furthermore, the features of the models are derived from tests routinely collected in the hospital. It can also serve as a general framework and allows integration of data from different hospitals and medical centers. These models can help to screen a large portion of negative cases at early stages of clinical diagnosis, prognosis, and treatment.

## 5. Conclusions

In this study, we showed that ML models can classify nonfunctional adrenal adenoma, subclinical CS, adrenal CS, and pituitary CS at screening test stage and follow-up test stage for the diagnosis of CS by utilizing selected input features derived from biochemical tests and imaging findings of patients who are suspected of CS.

We compared several prominent ML algorithms and demonstrated the ability of the RF-based models to accurately predict clinical interpretation of CS, despite the moderate size of the dataset and class imbalance problem. We think that success of the RF algorithm is because of its capabilities of handling small sample and imbalanced CS data, learning complex dependencies, reducing variance, inherently determining the cut-off levels of the features, and not requiring data scaling and standardization in advance.

Trained RF-based models on the T&V dataset and their predictive performance on FIT dataset show promising results. We suggest the use of ALLvsNF model to discriminate between NF and CS in the screening testing stage (Stage-1) for the diagnosis of CS in clinical evaluations. Furthermore, we suggest the use of multiclass ALL model to discriminate among subtypes of CS in the follow-up testing stage (Stage-2) for the identification of the cause of CS in clinical evaluations. Also, multiclass ALL model with Stage-1 features can be employed to get an early opinion about the subtype of CS for diagnosis, prognosis, and treatment choices.

The developed ML models outperformed the physician’s judgments under the constraint of using only the selected biochemical test findings utilized in ML model development. In Stage-1, physicians were able to classify subjects with F1 score of 76.6 % whereas F1 score of ML model was 98.5%. In Stage-2, the overall success rate of the physicians was low with an F1 score of 49.4% while ML model achieved an F1 score of 96.4%. These suggests that ML approach can help improve physician’s judgment in diagnosing CS subtypes with limited biochemical tests which are cumbersome and stressful for patients.

We provide a web application as clinical decision support tool for public use where a user can input patient’s test findings and receive prediction for CS.

Further work can be done to improve the performance of the proposed approach. We intend to update the models by adding new informative samples, adding new features derived from biochemical tests, and conduct experiments to further assess the clinical usefulness of the approach.

The overall results suggest that ML algorithms have potential to provide early detection and correct diagnosis of CS that may facilitate treatment and improve patient outcomes.

## Data Availability

The raw data is not available. However, authors made the mathematical models, raw results and software code available. A website for the proposed method is accessible publicly.

https://github.com/SenolIsci/cushing01

https://cushings-syndrome-prediction.herokuapp.com/.

## Acknowledgements

We thank Banu Erturk and Mehmet Eren Kalender for their expertise in clinical evaluations.

## Appendix A. Supplementary materials

Supplementary materials associated with this article can be found in the online version.

